# Seroprevalence, Knowledge, Attitudes, and Practices of People to COVID-19 in Chililabombwe and Lusaka Districts of Zambia

**DOI:** 10.1101/2025.09.08.25335374

**Authors:** Mercy Sampa, Joyce Siwila, Bernard Hangómbe, Gerald Misinzo, Mark Rweyemamu, Chisanga Chipanta, Martin C. Simuunza

## Abstract

Coronavirus disease 2019 (COVID-19) is a respiratory disease caused by the severe acute respiratory syndrome coronavirus 2 (SARS-CoV-2), which was first reported in Wuhan, China. Understanding seroprevalence and the knowledge, attitudes and practices (KAP) of individuals is a given context is crucial for effective public health interventions. This cross-sectional study assessed the seroprevalence of SARS-CoV-2 antibodies and KAP toward COVID-19 in Chililabombwe and Lusaka districts of Zambia. A total of 179 participants were enrolled in the study. These included individuals who visited a health facility, were 18 years of age or older, and were residents of the selected districts. Each participant was swabbed with one nasopharyngeal sample by the medical officer present and immediately subjected to a COVID-19 rapid diagnostic test using the Sure Status® kit (Premier Medical Corporation) targeting the nucleocapsid protein antigen from SARS-CoV-2. In addition, Samples were collected from these participants by drawing approximately 5 ml of venous blood, and the extracted sera was subjected to ELISA targeting the SARS-CoV-2 Nucleocapsid protein. A semi structured questionnaire was used to collect demographic and KAP data from the participants. Only one positive case was detected using the rapid diagnostic test (RDT) 0.5% (1/179). The overall seroprevalence of COVID-19 antibodies in the study areas was 9.5% (17/179). The seroprevalence in Chililabombwe (11.0%) was higher than that in Lusaka (8.0%). The study found variations in seroprevalence based on age, gender, and education level, with higher rates among individuals aged 40-49, females and those with no education. However, these differences were not statistically significant (*p* > 0.05). Both seropositive and seronegative participants exhibited moderate knowledge and attitudes, alongside high levels of preventive practices, indicating a shared understanding of health risks and practices. Further analysis showed that low knowledge, attitudes and practices were strongly associated with higher seropositivity. This study recommends enhanced community engagement in future pandemics to close knowledge gaps, disseminate accurate information, and promote effective preventive behaviours. These efforts could help limit the spread of diseases and strengthen community resilience in response to pandemics.

## Introduction

Coronavirus disease 2019 (COVID-19) is a respiratory disease caused by the severe acute respiratory syndrome coronavirus 2 (SARS-CoV-2) (Agrahari *et al*., 2021). The disease, (COVID-19), was first reported in Wuhan, China, in December 2019 (Khan, 2020). The principal route of transmission is either through inhalation of airborne droplets exhaled by infected individuals or direct contact of these droplets with the eyes, nose, or mouth (Wei *et al*., 2020). Through its spike (S) protein structure, the virus gains entry into the host by attaching to the angiotensin-converting enzyme 2 (ACE2) receptors on the surface of human cells. This facilitates its entry into host cells, particularly those in the respiratory system (Theodore *et al*., 2023). The virus causes similar symptoms to severe acute respiratory syndrome coronavirus (SARS-CoV) and Middle East respiratory syndrome (MERS-CoV) which usually begin to show two weeks after infection (Kontou, 2020). These include fever, respiratory problems or breathlessness, fatigue, muscle aches, reduced ability to taste or smell, headaches, and sensations of nausea or vomiting (Adam *et al*., 2022). However, COVID-19 disease has the potential to result in serious complications like kidney failure, severe pneumonia, acute respiratory syndrome, and fatality (Wei *et al*., 2020).

The diagnosis of COVID-19 includes molecular tests, antigen tests and serological tests. Low and middle-income countries usually utilize the antigen rapid diagnostic test (Ag RDT) because of its low cost. This is done through a nasopharyngeal swab (Jacobs *et al*., 2020). However, the most dependable method for detecting SARS-CoV-2 currently is via real-time reverse transcription polymerase chain reaction (RT-PCR). This technique pinpoints distinct genetic sequences of the virus, confirming its presence in samples collected from the upper respiratory tract (Mesina *et al*., 2021). Serological laboratory examinations identify SARS-CoV-2 antibodies within blood samples, these are quite simple to perform and easy to use for screening (Daka, 2014; Rubegwa, 2015).

The epidemiological landscape of COVID-19 in Zambia has been relatively well-defined (WHO, 2023). The Ministry of Health (MOH) reported the first COVID-19 case in March 2020, and as of August 17^th^, 2023, the country had seen approximately 349,287 confirmed cases and 4,069 deaths. (National Public Health Institute, 2022) However, given that Zambia still faces limitations in its testing capacity, the actual number of people exposed to the disease may be underestimated (Adam *et al*., 2022). The combined measured prevalence of SARS-CoV-2 was found to be 10.6% (95% CI of 7.3-13.9). The researchers concluded that the actual number of SARS-CoV-2 infections might surpass the officially recorded count in these six districts, emphasizing the importance of early isolation of infected individuals and timely identification of their contacts (Mulenga *et al*., 2021).

Various factors, including socioeconomic status, test kit availability, and proximity to the nearest healthcare facility, could impact an individual’s capacity to undergo testing. This can lead to underreporting of the prevalence of the disease in the country. However, the identification of anti-SARS-CoV-2 antibodies through serological assays may aid in approximating the number of individuals who may have been exposed to the infection.

Understanding the knowledge, attitudes, and practices (KAP) among individuals concerning COVID-19 can facilitate the development of targeted prevention and control strategies (Ugwu *et al*., 2020). Several studies have explored public KAP towards COVID-19 across different settings, highlighting how these factors influence the success of public health responses (e.g., Zhong et al., 2020; Reuben et al., 2021). In Zambia, however, there’s limited data linking KAP to actual exposure to the virus, especially in high-traffic or border areas. The main goal of this study was to determine the seroprevalence of antibodies to SARS-CoV-2 in adults and investigate their knowledge, attitudes, and practices of COVID-19 in Chililabombwe and Lusaka district of Zambia.

## Materials and Methods

### Study Area

The study was conducted among residents of Lusaka and Chililabombwe districts in Zambia (Figure 1). These districts were purposefully selected due to their strategic roles as transit hubs and their geographical location within provinces reporting the highest cumulative COVID-19 cases in the country (Ministry of Health, 2023). Lusaka and Chililabombwe districts are both urban areas with populations of 2,567,093 and 124,577, respectively (Central Statistics Agency, 2022). Chililabombwe, a major border town with the Democratic Republic of Congo, hosts one of Zambia’s largest cross-border markets. Meanwhile, Lusaka is home to Kenneth Kaunda International Airport, the country’s largest and busiest airport. According to Ministry of Health data from 2023, Lusaka Province recorded a total of 94,242 COVID-19 cases, while Copperbelt Province reported 47,795 cases out of the 349,287 at country total.

**Figure 1.**
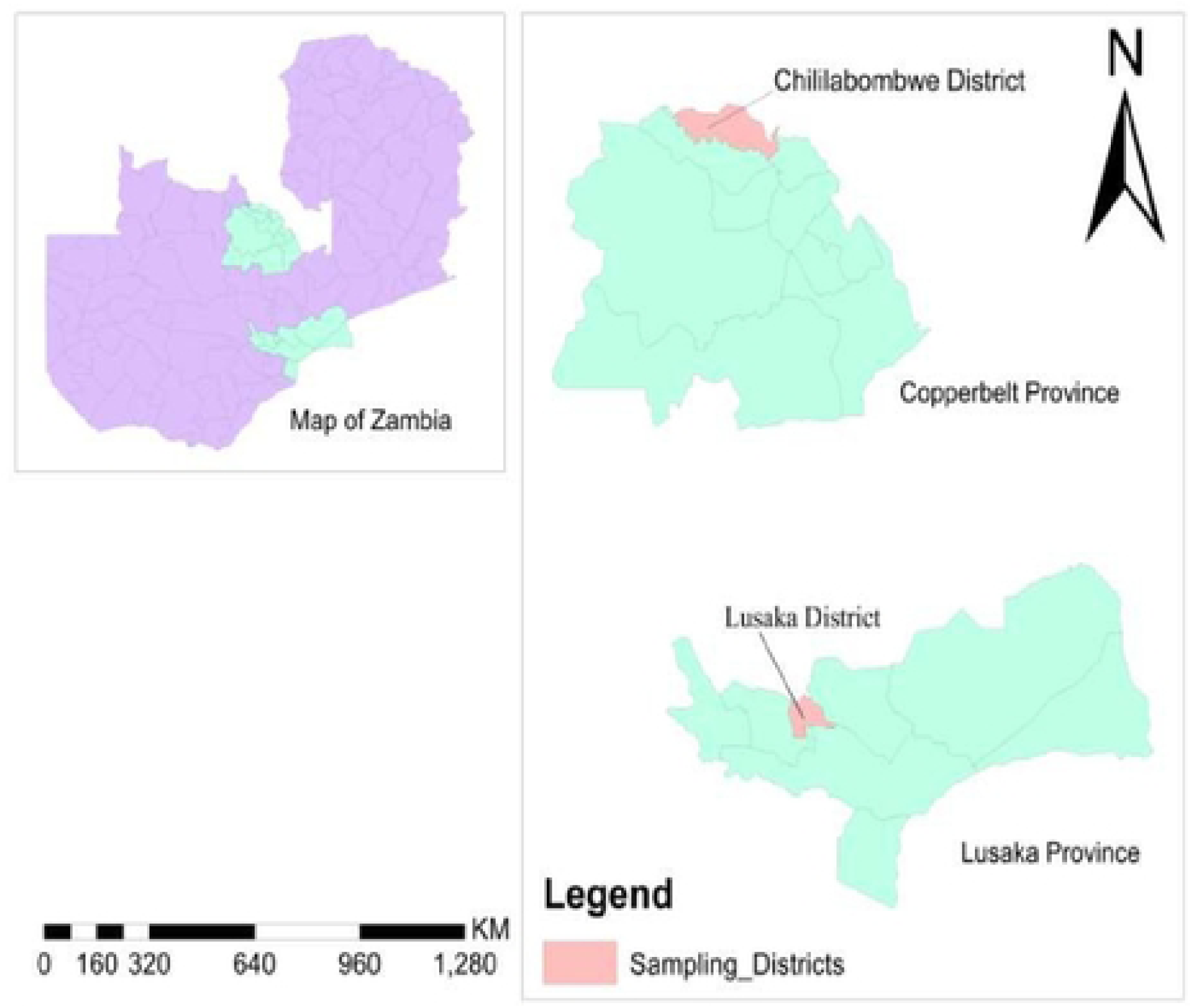
Map of the Study Area

### Ethics approval and consent to participate

The study was approved by Excellence in Research Ethics and Science (ERES) CONVERGE IRB (Reg No.2022-June-014) and the National Health Research Authority (NHRA) (Reg No. 20 NHRA000003/19/01/2023). Further permission was sought from the Ministry of Health and authorization to enter health facilities was obtained from the Provincial Health Office, District Health Offices and personnel in-charge of each health facility. Informed consent was sought from all those that agreed to participate in the study and information collected from the participants was used only for research purposes. Confidentiality of personal information was upheld throughout the study by ensuring that participants’ information was anonymous and identities withheld.

### Study design and sampling

This study was cross-sectional in design. The sample size was determined using the formula recommended by Charan and Biwas (2013) assuming a SARS-CoV-2 prevalence of 13.5% in Zambia (Shanaube *et al*., 2022), a 95% confidence level and a 5% absolute error. The resulting sample size calculation was estimated to be 179 participants. This study included individuals who visited a health facility, were 18 years of age or older, residing in the district of interest, were willing, and provided consent to participate in the research. Each participant was swabbed with one nasopharyngeal sample by the medical officer present and immediately subjected to a COVID-19 rapid diagnostic test using the Sure Status® kit (Premier Medical Corporation) targeting the nucleocapsid protein antigen from SARS-CoV-2. In addition, blood samples were collected from the participants by drawing approximately 5 ml of venous blood samples from either the median cubital or cephalic vein and placed into plain vacutainers. After sample collection, a structured questionnaire was administered to each participant to collect information on demographics, COVID-19 symptoms, household COVID-19 cases, perception of COVID-19 infection risk; adherence to recommended prevention measure, knowledge, attitudes and practices regarding COVID-19.

### Sample processing and analysis

The collected blood samples were left overnight and centrifuged the following day at 1500 rpm for 10 minutes. The resulting serum was harvested and stored at −20°C in compliance with cold-chain protocols. They were subsequently transported to the University of Zambia, School of Veterinary Medicine Public Health laboratory on ice. The Invitrogen™ Human SARS-CoV-2 N Enzyme-linked Immuno-Sorbent Assay (ELISA) (Thermo Fisher catalogue number EH490 RB) targeting the nucleocapsid protein was used to detect antibodies to SARS-Cov-2, according to the manufacturer’s instructions. Optical density (OD) readings were taken at 450 nm using a Multiskan™ FC Microplate Photometer (Thermo Fisher Scientific, USA). Sample results were interpreted based on the manufacturer’s recommended cut-off values: samples with OD values above the cut-off were considered positive, while those below were classified as negative.

### Data analysis

Data were entered into Microsoft Excel and statistical analyses were conducted using IBM SPSS Statistics version 26 (SPSS Inc., Chicago IL, USA) and GraphPad Prism 9.0 (GraphPad Software, Boston, Massachusetts USA). Seroprevalence was determined as the proportion of individuals with a positive test result for the total antibodies. The Pearson Chi-squared (*X^2^*) test or the Fisher’s Exact test, where appropriate were used to examine associations between sociodemographic factors and seropositivity.

The participants’ KAP were assessed by summing the correct responses, from which the mean score was calculated. Participants responded with “Yes” or “No,” with correct answers scoring 1 and incorrect answers scoring 0. The KAP levels were categorised as high, moderate, or low based on predetermined thresholds using Bloom’s cut-off criteria (Hasan et al., 2022) According to these criteria, scores above 60 indicated a high level of KAP, scores between 25 and 59 represent moderate levels, and scores below 25 suggest a low level. All statistics were considered significant at a *p* ≤ 0.050.

## Results

### Characteristics of study participants and seroprevalence of SARS-CoV-2

Of the 179 participants, 88 (49.2%) were male (Table 1). A total of 84 respondents (46.9%) were aged between 18 and 29 years, representing the largest age group in the study. The smallest age group comprised individuals above 50 years, with 15 respondents (8.4%) in this category. In terms of marital status, the majority (57.5%) were single, followed by 66 participants (36.9%) who reported being married.

**Table 1.** Socio-demoRraphic characteristics and seroprevalence study participants

| Variable | Categories | No. | No. positive (%; 95% CI) | p-value |
| --- | --- | --- | --- | --- |
| Overall |  | 179 | 17 (9.5, 6.0-14.7) |  |
| District | Chililabombwe | 91 | 10 (11.0, 6.1-19.1) | 0.49 |
|  | Lusaka | 88 | 7 (8.0, 3.9-15.5) |  |
| Gender | Male | 84 | 5 (6.0, 2.6-13.2) | 0.13 |
|  | Female | 95 | 12 (12.6, 7.4-20.8) |  |
| Age group (Years) | 18 – 29 | 94 | 10 (10.6, 5.9-18.5) | 0.71 |
|  | 30 – 39 | 48 | 3 (6.2, 2.1-16.8) |  |
|  | 40 - 49 | 22 | 3 (13.6, 4.7-33.3) |  |
|  | Above 50 | 15 | 1 (6.7, 1.2-29.8) |  |
| Marital status | Married | 75 | 8 (10.7, 5.5-19.7) | 0.12 |
|  | Single | 102 | 8 (7.8, 4.0-14.7) |  |
|  | Widow/widower | 2 | 1 (50.0, 9.5-90.5) |  |
| Level of Education | College/University | 60 | 2 (3.3, 0.9-11.4) | 0.16 |
|  | Secondary | 73 | 9 (12.3, 6.6-21.8) |  |
|  | Primary | 37 | 4 (10.8, 4.3-24.7) |  |
|  | No education | 9 | 2 (22.2, 6.3-54.7) |  |
| Employment status | Employed | 93 | 9 (9.7, 5.2-17.4) | 0.93 |
|  | Unemployed | 86 | 8 (9.3, 4.8-17.3) |  |

The majority (53.6%) of the respondents reported secondary education as the highest level of education they had attained while 5.6% of the respondents had no formal education. About 52.0% were employed, and participants were evenly distributed between the two districts, with 50.8% from Chililabombwe district (Table 1).

Only one positive case was detected using the rapid diagnostic test (RDT). However, ELISA testing, revealed an overall SARS-CoV-2 antibody seroprevalence of 9.5% (95% CI: 6.0–14.7) (Table 1). Chililabombwe recorded a higher seroprevalence of 11.0% (95% CI: 6.1–19.1), compared to 8.0% (95% CI: 3.9–15.5) in Lusaka. Variations in SARS-CoV-2 seropositivity were observed across demographic variables such as age group, sex, marital status, education level, and employment status (Table 1). Apart from lack of formal education, no other demographic variable showed a statistically significant association with SARS-CoV-2 seropositivity in the two districts (Table 1).

### Knowledge, attitudes and practices of study participants and seroprevalence of COVID-19

Out of the 179 respondents (both seropositive and seronegative for COVID-19), 95.5% reported that they had heard of COVID-19. Among those who were aware, 9.4% were seropositive, while 90.6% were seronegative. The remaining 4.5% had no prior awareness of the disease.The source of information regarding COVID-19 was investigated in this study. Respondents from both the seropositive and seronegative groups indicated that they received information about COVID-19 through various sources (Figure 4), with the majority relying on friends, family, and colleagues. Other sources included TV/Radio, social media, health workers, newspapers, religious leaders, and teachers. Both seropositive and seronegative participants identified the common COVID-19 symptoms, including cough, runny nose, and fever. They understood close contact as the main mode of transmission, though fewer recognized airborne droplets. Most believed that COVID-19 was curable, mainly by seeking help from healthcare facilities, while a few mentioned herbal remedies or prayer. Overall, both seropositive and seronegative groups showed moderate knowledge of COVID-19 (Table 3).

**Figure 4.**
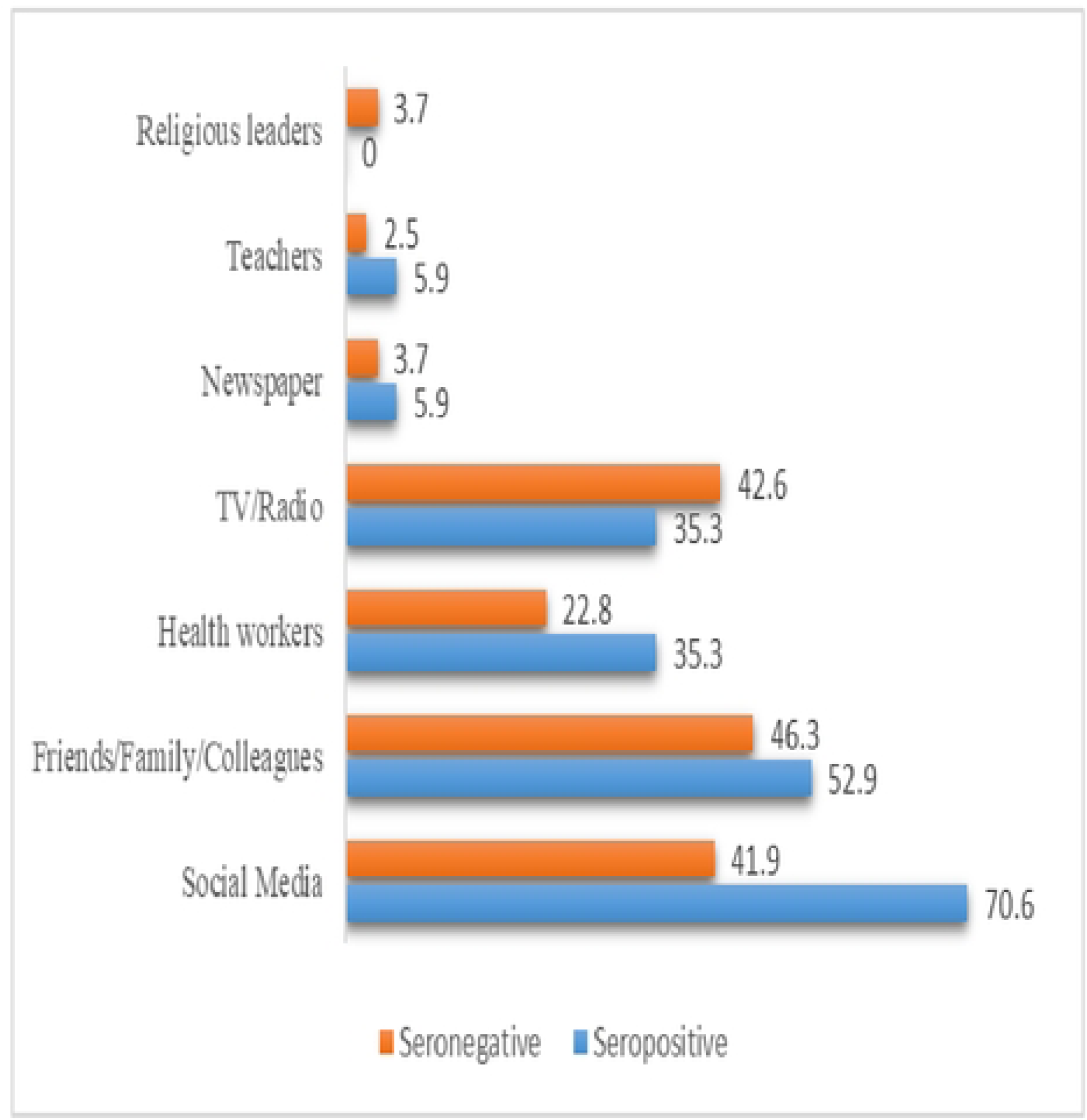
Participant’s source of information on COVID-19. The number of respondents for seropositive and seronegative were 17 and 162 respectively.

**Table 2.**
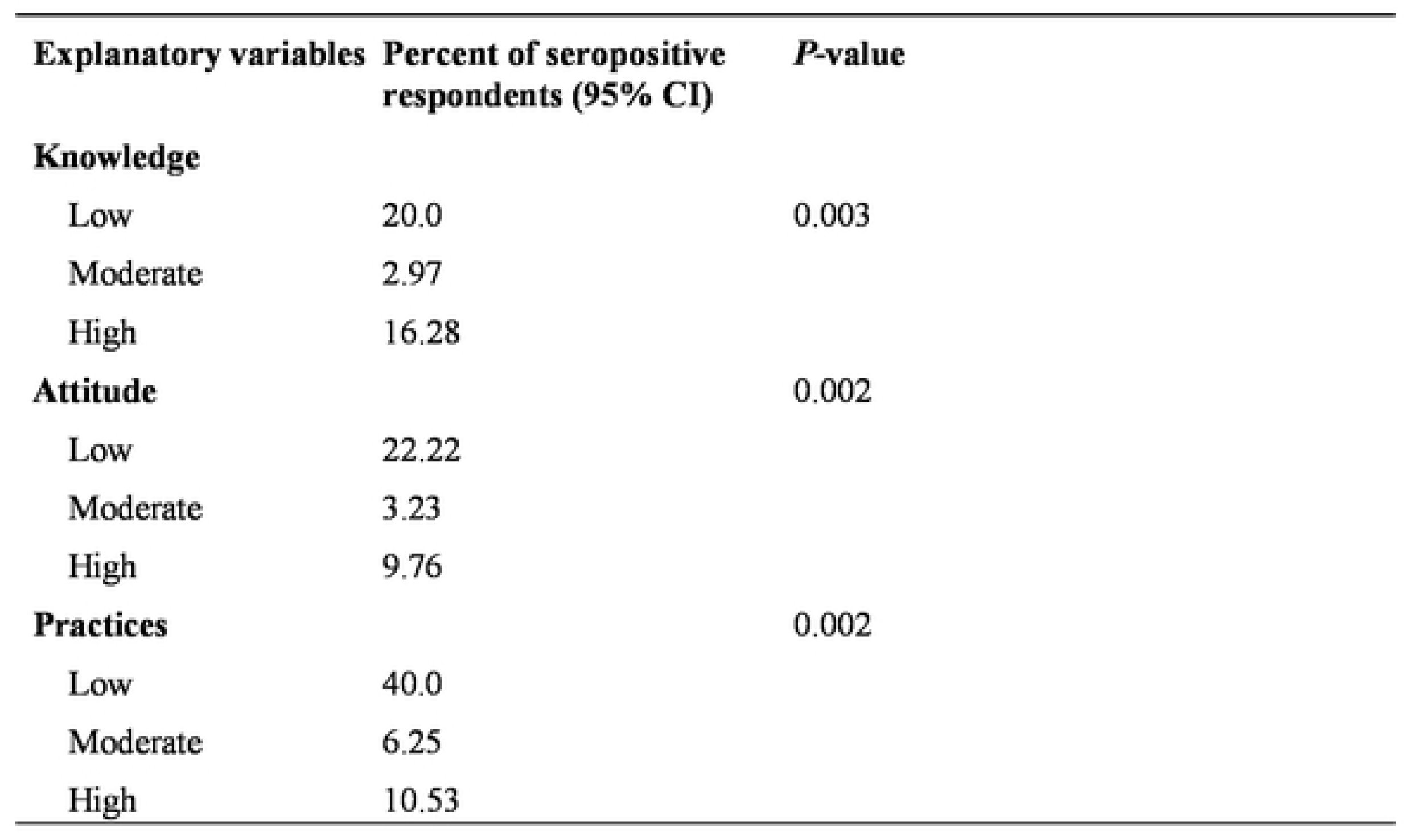
Association of Knowledge, Attitude, and Practices with SARS-CoV-2 positivity.

**Table 3.** Participants knowledge on COVID-I9

| Variable | Levels | Frequency<br>seropositive | of<br>Proportion<br>(%) | Score<br>(%) | Frequency<br>seronegative | Of<br>Proportion<br>(%) | Score<br>(%) |
| --- | --- | --- | --- | --- | --- | --- | --- |
| **What is the causative agent for COVID-19?** | Bacteria | 8 | 47.05 |  | 27 | 16.7 |  |
|  | Virus | 9 | 52.94 | 52.94* | 135 | 83.33* | 83.33 |
| **Signs/ symptoms of COVID-19** | Coughing | 6 | 35.3 |  | 81 | 50 |  |
|  | Runny nose | 9 | 52.9 | 30.78* | 73 | 45.1 | 40.38* |
|  | Headache | 1 | 5.9 |  | 53 | 32.7 |  |
|  | Fatigue | 2 | 12.8 |  | 23 | 14.2 |  |
|  | Fever | 8 | 47 |  | 97 | 59.9 |  |
| **How can a person get COVID-19?** | Through handshake | 6 | 35.2 |  | 53 | 32.7 | 30.03* |
|  | Air droplets | 3 | 17.6 |  | 47 | 29 |  |
|  | Close contact with infected person | 7 | 41.2 | 31.3* | 46 | 28.4 |  |
| **Do you think COVID-19 can be cured?** | Yes | 15 | 88.2 | 88.2* | 123 | 75.9 | 75.9* |
|  | No | 2 | 12.8 |  | 39 | 24.1 |  |
| **How can COVID-19 be cured?** | Herbal remedies | 0 | 0 |  | 14 | 8.6 |  |
|  | Praying | 1 | 5.9 |  | 2 | 1.2 |  |
|  | By health personal | 13 | 76.5 | 76.5* | 117 | 72.2 | 72.2* |
| **Average score on knowledge of COVID-19** |  |  |  | 55.94** |  |  | 52.68** |

The study found that participants who had low knowledge about COVID-19 had the highest seropositivity 20% followed by those with high knowledge (16.3%) and moderate knowledge (2.9%) (Table 2). The association between knowledge levels and seropositivity was statistically significant (p = 0.003) (Table 2).

Seropositive individuals with low attitude scores had the highest seropositivity rate at (22.2%), compared to (9.8%) among those with high attitude scores and (3.2%) for moderate attitudes. (Table 2). This trend suggested that individuals with a poorer attitude towards COVID-19 prevention measures were more likely to have been exposed to the virus.

A Similar pattern was found in practice scores: participants with low adherence to preventive practices had a seroprevalence of (40%), compared to (10.5%) for those with high practice levels and (6.3%) for moderate (Table 2). This pattern indicated variations in infection rates based on the level of preventive practices adopted. There was a significant association between individuals’ practices and their likelihood of testing positive for SARS-CoV-2 (p = 0.002).

Despite similar emotional responses to COVID-19 both those who tested seropositive and those who tested seronegative for SARS-CoV-2 believed they could get COVID-19 including embarrassment, fear, and sadness (Table 4). Both groups primarily feared death and isolation, and most seropositive participants sought emotional support from doctors or family

**Table 4.** Participants attitude towards C0VID-19

| Variable | Levels | Frequency<br>seropositive | of<br>Proportion<br>(%) | Score<br>(%) | Frequency<br>seronegative | of<br>Proportion (%) | Score (%) |
| --- | --- | --- | --- | --- | --- | --- | --- |
| **Do you think**<br>**you can get**<br>**COVID-19?** | Yes | 13 | 76.47 | 76.47* | 132 | 81.48 | 81.48* |
|  | No | 0 | 0 |  | 27 | 16.7 |  |
|  | Maybe | 4 | 23.53 |  | 3 | 1.85 |  |
| **Do you know**<br>**anyone in**<br>**your**<br>**community**<br>**who had**<br>**COVID-19?** | Yes | 6 | 35.29 | 35.29* | 78 | 48.15 | 48.15* |
|  | No | 11 | 64.71* |  | 84 | 51.85 |  |
| **How would**<br>**you feel if you**<br>**were found**<br>**that you have**<br>**COVID-19?** | Embarrassed | 1 | 5.88 | 20.0* | 3 | 1.85 | 26.42* |
|  | Fear | 3 | 17.65 |  | 52 | 32.09 |  |
|  | Surprised | 1 | 5.88 |  | 7 | 4.32 |  |
|  | Sadness | 5 | 29.41 |  | 36 | 22.22 |  |
|  | Other | 7 | 41.17 |  | 64 | 39.51 |  |
| **Who would**<br>**you talk to if**<br>**you had**<br>**COVID-19?** | Parent | 6 | 35.29 | 27.45* | 33 | 20.37 | 21.91* |
|  | Children | 3 | 17.64 |  | 28 | 17.28 |  |
|  | Doctor | 5 | 29.41 |  | 57 | 35.19 |  |
|  | Spouse | 0 | 0 |  | 24 | 14.81 |  |
|  | No one | 3 | 17.65 |  | 20 | 12.35 |  |
| **What worries**<br>**you the most**<br>**when you**<br>**think of**<br>**COVID-19?** | Isolation | 3 | 17.65 | 50* | 29 | 17.90 | 31.48* |
|  | Unemployment | 0 | 0 |  | 11 | 6.79 |  |
|  | Death | 14 | 82.35 |  | 113 | 69.75 |  |
|  | Other | 0 | 0 |  | 9 | 5.55 |  |
| **Average KAP**<br>**score on**<br>**Attitude of**<br>**COVID-19** |  |  |  | 41.84** |  |  | 41.89** |

The findings in Table 5 indicate that a majority of respondents consistently followed mask-wearing guidelines in public places. Most (52.94%) of the participants followed hand-washing practices. Furthermore, an overwhelming majority (88.2%) consistently avoided large gatherings. The average scores indicated high adherence to COVID-19 preventive practices in both the seropositive and seronegative participants (Table 5).

**Table 5.** Participants Practices towards C0V1D-19

| Variable | Levels | Frequency<br>seropositive | of<br>Proportion<br>(%) | Score<br>(%) | Frequency<br>seronegative | of<br>Proportion<br>(%) | Score<br>(%) |
| --- | --- | --- | --- | --- | --- | --- | --- |
| **Have you consistently followed the recommended guidelines for wearing masks in public places?** | Yes | 13 | 76.47 | 76.47* | 116 | 71.60 | 71.60* |
|  | No | 0 | 0 |  | 1 | 0.62 |  |
|  | Sometimes | 4 | 23.53 |  | 45 | 27.78 |  |
| **How frequently do you wash your hands with soap and water as a preventive measure for COVID-19?** | Always | 9 | 52.94 | 52.94* | 96 | 59.26 | 59.26* |
|  | Frequently | 2 | 11.76 |  | 22 | 13.58 |  |
|  | Occasionally | 6 | 35.29 |  | 44 | 27.16 |  |
| **In the past month, how often have you avoided large gatherings or crowded places to reduce the risk of COVID-19 transmission?** | Always | 15 | 88.24 | 88.24* | 135 | 83.33 | 83.33* |
|  | Frequently |  | 0 |  | 23 | 14.19 |  |
|  | Occasionally | 2 | 11.76 |  | 4 | 2.47 |  |
| **Do you think we can prevent such a global pandemic in the future?** | Yes | 10 | 58.82 | 58.82* | 103 | 63.58 | 63.58* |
|  | No | 0 | 0 |  | 9 | 5.55 |  |
|  | Maybe | 7 | 41.18 |  | 50 | 30.86 |  |
| **Average score on Practices of COVID-19** |  |  |  | 69.12** |  |  | 69.44** |

% = Percentage; \* = Proportion considered as practice score, \*\*=Average practice score

## Discussion

Coronavirus disease 2019 (COVID-19) has impacted countries globally, with varying degrees of disease burden and social economic disruptions, prompting various levels of responses to the disease. This study aimed to estimate the prevalence of SARS-CoV-2 antibodies using both rapid diagnostic tests (RDTs) and ELISA. While the RDT detected only one positive case out of 179 participants, ELISA revealed a seroprevalence of 9.5%. The prevalence from the RDT was much lower than that of ELISA because RDTs detected active infection by identifying viral antigens and had lower sensitivity, especially in individuals with low viral loads or during the later stages of infection (Mak et al., 2020; Peeling et al., 2021). ELISA, on the other hand, detected a higher number of participants who were positive to SARS-CoV 2 antibodies, indicating prior exposure, making it more reliable for assessing cumulative exposure in the population. The markedly lower prevalence detected by RDTs compared to ELISA could be attributed to their lower sensitivity in detecting active infection, particularly among asymptomatic individuals, those with low viral loads, or in the later stages of infection as the antigen levels declined (Dinnes et al., 2021; Peeling et al., 2021). In contrast, ELISA detects antibodies that persist after infection, making it a more reliable method for assessing cumulative exposure in the population. Similar findings were reported in studies from

Mozambique (Arnaldo et al., 2022), Nigeria (Reuben et al., 2021), and Ethiopia (Aynalem et al., 2021), where serological assays demonstrated significantly higher prevalence rates than RDTs, highlighting the limitations of relying solely on antigen-based tests for accurate population-level surveillance in African settings.

Chililabombwe recorded a slightly higher seropositivity than Lusaka, a difference that may be attributed to factors such as socioeconomic conditions, cross border activities, healthcare infrastructure, and access to testing services. Similar disparities in seroprevalence have been reported elsewhere. For instance, studies in Mozambique and the United States of America linked regional variations in infection rates to differences in healthcare infrastructure and socioeconomic status (Naiyer et al., 2021; Hatef et al., 2020).

A higher prevalence was observed among individuals aged 18-29, suggesting that younger adults may have been more socially active, thus increasing their risk of exposure. This aligns with research in Mozambique, where older adults (≥55 years) showed greater adherence to preventive measures due to perceived vulnerability (Arnaldo et al., 2022). Although females had higher seroprevalence than males in this study, the difference was not statistically significant. This finding is consistent with a systematic review in Africa, which found no clear link between gender and antibody presence (Chisale et al., 2021), and the WHO’s neutral stance on gender in COVID-19 health planning (WHO, 2022).

Among all assessed factors, lack of formal education and moderate attitude levels were significantly associated with higher seropositivity. Individuals with no formal education may have limited health literacy, hindering their ability to understand or act on public health information (Smith et al., 2020 Interestingly, individuals with moderate attitude scores demonstrated lower seropositivity than those with low scores. This pattern may indicate that a moderate level of concern—striking a balance between complacency and alarm—could be linked to more consistent adherence to protective behaviours. However, further investigation is warranted to better understand this relationship (Jones et al., 2021).

In line with previous studies (Zhong et al., 2020; Aynalem et al., 2021), this study found that low knowledge, low attitude, and low practice scores were associated with increased risk of SARS-CoV-2 exposure. Participants with low KAP scores were more likely to test positive, reinforcing the need for targeted health education and behavioural interventions to improve knowledge, attitudes, and practices within the community. Participants demonstrated a reasonable understanding of COVID-19’s cause, symptoms, and primary transmission methods. These findings mirror earlier research (Zhong et al., 2020; Clements, 2020), which attributed public awareness to effective health communication. However, misconceptions remained. While most recognized close contact as a key transmission route, fewer participants identified handshakes and airborne droplets—consistent with findings by Geldsetzer (2020). Although many believed COVID-19 could be cured, often relying on healthcare providers, few cited herbal remedies or prayer. This reliance on medical professionals reflects public trust in health systems, even amid uncertainty.

Information about COVID-19 was accessed through diverse sources, including health workers, family, friends, radio/TV, religious leaders, and newspapers. This highlights the importance of leveraging multiple communication channels in public health messaging. Health professionals, in particular, were viewed as trusted figures (Schiavo, 2014), underscoring their central role in effective information dissemination. Likewise, religious leaders and health educators were noted as influential sources due to their embedded roles in communities (Berkley, 2020; Basch et al., 2020).

The study also found that many participants acknowledged their susceptibility to COVID-19, expressing a range of emotions such as fear, sadness, and concern, similar to findings by Geldsetzer (2020) and Reuben et al. (2021). Despite this emotional response, the moderate average attitude scores suggest a general acceptance of the disease’s presence and seriousness in the communities studied.

Preventive practices were generally well adhered to. Most participants reported consistently wearing face masks in public, a trend supported by global evidence (Chu et al., 2020). Social distancing was also widely practiced. However, handwashing compliance was less consistent, echoing previous findings that, despite widespread promotion, hand hygiene practices often fall short (Luby et al., 2021). The overall high practice scores in this study reflect a strong community commitment to following recommended health measures.

We acknowledge the limitations of the study. The serological tests were conducted towards the end of the pandemic and may have yielded different results compared to those performed at the peak of the outbreak. A larger sample size may have given a broader perspective on social variables such as employment status and other social factors investigated.

## Conclusions

The prevalence of SARS-CoV-2 antibodies in Chililabombwe and Lusaka districts was estimated to be 11% and 8%, respectively. Both seropositive and seronegative participants demonstrated moderate knowledge and attitudes, as well as high levels of practice, which suggests a shared understanding of health risks and preventive measures of COVID-19. Lower knowledge, low attitudes and low practices were strongly associated with higher seropositivity. There is need to develop systems that incorporate various variables like climatic change and variability to enhance community engagement in future pandemics while monitoring the prevalence and dynamics of COVID-19 continuously.

## Data Availability

All data generated or analyzed during this study are included in this published article and its supplementary information files. Additional datasets used and/or analyzed during the current study are available from the corresponding author on reasonable request. Data containing sensitive participant information are available under controlled access to protect participant confidentiality, in accordance with ethical approvals from ERES CONVERGE IRB and the National Health Research Authority (NHRA).

## Acknowledgements

We would like to express our sincere gratitude to SACIDS – Africa Centre of Excellence for Infectious Diseases, Sokoine University of Agriculture, Morogoro, Tanzania, for the invaluable support. Special thanks also go to Dr. Chibeza Zulu, Mr. Andrew Mukubesa, Mr. Mwelwa Chembensofu, Mr. Patrick Katemangwe, and Mr. Penjaninge Kapila for their consistent technical assistance and collaborative brainstorming sessions. We are also grateful to the Mr Japhet Chiwaula of National Malaria Elimination Centre and the Ministry of Health for providing the essential facilities that greatly supported the data collection process.

## References

1. Adam M, Mohamoud J, Mohamood A, Mohamed A, Garba B, Dirie N. Seroprevalence of Anti-SARS-CoV-2 Antibodies in Bendir Region, Somalia. Vaccines. 2022;:1–11.

2. Agrahari R, Mohanty S, Vishwakarma K, Nayak SK, Samantaray D, Mohapatra S. Update vision on COVID-19: Structure, immune pathogenesis, treatment and safety assessment. Sensors Int. 2021;:1–9.

3. Al-Tawfiq JA, Al-Homoud AH, Memish ZA. Hand hygiene in the COVID-19 era. J Infect Public Health. 2021;14(5):665–6.

4. Arnaldo P, Mabunda N, Young WP, Tran T, Sitoe N, Chelene I, Ismeal N. Mozambican Population: A Cross-Sectional Serologic Study in 3 Cities, July–August 2020. Clin Infect Dis. 2022;:S285–93.

5. Aynalem YA, Yirgu R, Gebresilassie M. Knowledge, Attitudes, and Practices towards COVID-19 and associated factors among healthcare workers in Southern Ethiopia. J Public Health Res. 2021;10(1):123–30.

6. Ball-Rokeach SJ, Kim YC, Matei S. Storytelling neighborhood: Paths to belonging in diverse urban environments. Commun Res. 2001;28(4):392–428.

7. Basch CE, Basch CH, Hillyer GC, Jaime C. The role of schools in promoting health literacy in the context of COVID-19. Prev Chronic Dis. 2020;17:E53.

8. Chu DK, Akl EA, Duda S, Solo K, Yaacoub S, Schünemann HJ. Physical distancing, face masks, and eye protection to prevent person-to-person transmission of SARS-CoV-2 and COVID-19: a systematic review and meta-analysis. Lancet. 2020;395(10242):1973–87.

9. Clements JM. Knowledge and behaviors toward COVID-19 among US residents during the early days of the pandemic: Cross-sectional online questionnaire. JMIR Public Health Surveill. 2020;6(2):e19161. doi:10.2196/19161.

10. Daka V. Seroprevalence and risk factors of toxoplasmosis in individuals attending Chipokotamayamba Clinic in Ndola, Zambia. 2014.

11. Dinnes J, Deeks JJ, Berhane S, Taylor M, Adriano A, Davenport C, Dittrich S, Emperador D, Takwoingi Y, Cunningham J, Beese S. Rapid, point-of-care antigen and molecular-based tests for diagnosis of SARS-CoV-2 infection. Cochrane Database Syst Rev. 2021;3. doi:10.1002/14651858.CD013705.pub2.

12. Geldsetzer P. Knowledge and perceptions of COVID-19 among the general public in the United States and the United Kingdom: A cross-sectional online survey. Ann Intern Med. 2020;173(2):157–60. doi:10.7326/M20-0912.

13. Hasan NIA, Abidin SZ, Ganggayah MD, Jamal NF, Aziz WNHWA. Knowledge, attitude and practices (KAP) theory towards preventive measures among Malaysians in early outbreak of COVID-19. Malays J Public Health Med. 2022;22(1):38–47.

14. Jacobs J, Kühne V, Lunguya O, Affolabi D, Hardy L, Vandenberg O. Implementing COVID-19 (SARS-CoV-2) rapid diagnostic tests in Sub-Saharan Africa: a review. Front Med. 2020;7:557797.

15. Jones M, Green R, White S. Attitudinal determinants of compliance with public health guidelines during the COVID-19 pandemic. Health Psychol Rev [Preprint]. 2021.

16. Khan MA. COVID-19: A global challenge with old history, epidemiology and progress so far. Molecules. 2020;1(26).

17. Kontou PI. Antibody tests in detecting SARS-CoV-2 infection: A meta-analysis. Diagnostics. 2020;:1–15.

18. Luby SP, Agboatwalla M, Feikin DR, Painter J, Billhimer W, Altaf A, Hoekstra RM. Effect of handwashing on child health: A randomised controlled trial. Lancet. 2021;366(9481):225–33.

19. Mak GC, Cheng PK, Lau SS, Wong KK, Lau CS, Lim WW. Evaluation of rapid antigen test for detection of SARS-CoV-2 virus. J Clin Virol. 2020;129:104500. doi:10.1016/j.jcv.2020.104500.

20. Mesina F, Mangahas C, Gatchalian EM, Ramos M, Torres R, Ariola S. Use of convalescent plasma therapy among hospitalized coronavirus disease 2019 (COVID-19) patients: A single-center experience. MedRxiv. 2021;:1–22.

21. Peeling RW, Olliaro P, Boeras DI, Fongwen N. Scaling up COVID-19 rapid antigen tests: promises and challenges. Lancet Infect Dis. 2021;21(9):e290–5. doi:10.1016/S1473-3099(21)00048-7.

22. Peiris JSM, Lai ST, Poon LLM, Guan Y, Yam LYC, Lim W, Nicholls J, Yee WKS, Yan WW, Cheung MT, Cheng VCC, Chan KH, Tsang DNC, Yung RWH, Ng TK, Yuen KY. Coronavirus as a possible cause of severe acute respiratory syndrome. Lancet. 2003;361(9366):1319–25. doi:10.1016/S0140-6736(03)13077-2.

23. Reuben RC, Danladi MMA, Saleh DA, Ejembi PE. Knowledge, attitudes, and practices towards COVID-19: An epidemiological study in North-Central Nigeria. J Community Health [Preprint]. 2021;45(5):1091–8. doi:10.1007/s10900-020-00881-1.

24. Rubegwa BA. Seroprevalence and risk factors of bovine brucellosis in dairy and traditional cattle herds in Kibaha District of Tanzania [thesis]. 2015;1.

25. Schiavo R. Health communication: from theory to practice. 2nd ed. San Francisco: Jossey-Bass; 2014.

26. Smith A, Jones B, Thompson C. The impact of educational attainment on health literacy and its role in the prevention of COVID-19. Public Health Rev [Preprint]. 2020.

27. Theodore D, Branche A, … LZ-J network, U. Clinical and demographic factors associated with COVID-19, severe COVID-19, and SARS-CoV-2 infection in adults: a secondary cross-protocol analysis of 4. JAMA Netw Open. 2023;:1–11.

28. Ugwu C, Adekola A, Fasoro O, Oyesola O, Heeney J, Happi C. Insights into the Nigerian COVID-19 outbreak. Semantic Scholar. 2020;:1–9.

29. Wei L, Zhang B, Lu J, Liu S, Chang Z, Peng C, Liu X. Characteristics of household transmission of COVID-19. Clin Infect Dis. 2020;:1943–6.

30. Zambia Statistics Agency. The Census of Population and Housing. 2022. Available from: https://www.zamstats.gov.zm/ (Accessed 12 Dec 2023).

31. Zhong BL, Luo W, Li HM, Zhang QQ, Liu XG, Li WT, Li Y. Knowledge, attitudes, and practices towards COVID-19 among Chinese residents during the rapid rise period of the COVID-19 outbreak: a quick online cross-sectional survey. Int J Biol Sci. 2020;16(10):1745– 52.

